# Effect of environmental factors in reducing the prevalence of schistosomiasis in schoolchildren: A panel analysis of three extensive national prevalence surveys in Brazil (1950–2018)

**DOI:** 10.1101/2022.09.12.22279843

**Authors:** Mariana Cristina Silva Santos, Guilherme Lopes de Oliveira, Sueli Aparecida Mingoti, Léo Heller

**Affiliations:** Instituto René Rachou, Fiocruz Minas, Belo Horizonte, Minas Gerais, Brasil.; Departamento de Computação, Centro Federal de Educação Tecnológica de Minas Gerais.; Departamento de Estatística, Universidade Federal de Minas Gerais, Brasil.

**Keywords:** Schistosomiasis mansoni, *Schistosoma* infection, water supply, sewage, ecological studies, Brazil

## Abstract

**Background:** Over seven decades, Brazil has made admirable progress in controlling schistosomiasis, and a frequent question about the explanation for this reduction refers to the effect of improving environmental factors in the country. This article seeks to identify factors related to the change in the epidemiological situation of schistosomiasis mansoni infection by analyzing three national prevalence surveys conducted since 1950.

**Methodology/principal findings:** This is an ecological study analyzing an unbalanced panel of data based on national surveys and considering the municipality as the unit of analysis. The sample consisted of 1,721 Brazilian municipalities, in which a total of 1,182,339 schoolchildren aged 7–14 was examined during the three periods corresponding to each survey (1947–1952, 1975–1979, and 2011–2015). Zero-inflated Poisson regression models, with mixed and random effects, were adjusted to assess the association between candidate factors and disease prevalence using a significance level of 5%. There was a significant decrease in disease prevalence between the first and last periods analyzed (RR 0.214, CI 0.184 – 0.249), with a protective association with access to sanitation (RR 0.996, CI 0.994 – 0.998), urbanization (RR 0.991, CI 0.989 – 0.993), and living in own households (RR 0.986, CI 0.983 – 0.989); and an inverse association with the water supply (RR 1.010, CI 1.008 – 1.011).

**Conclusion:** The findings of this study indicate a decrease in the prevalence of schistosomiasis over seven decades in schoolchildren from the analyzed Brazilian municipalities, mediated by environmental factors and social conditions. The increased access to piped water in the municipalities apparently triggers other ways of contact with watercourses, generating new transmission routes and suggesting the need for a systemic approach concerning contact with water.

**Author Summary:** Schistosomiasis mansoni is a neglected tropical disease caused by infection from parasitic worms of the species *Schistosoma mansoni*. Due to the complexity of the mechanism of transmission and maintenance of schistosomiasis, several preventive actions on diverse conditioning factors can promote disease control. Active search, timely treatment of cases, stool tests, and epidemiological investigations are the initial actions under programs for epidemiological surveillance of the disease. Thus, momentum historical landmark surveys on the national prevalence of the disease can provide valuable information about its epidemiological pattern over the years. Our study addressed three national surveys with historical coverage (1950, 1970, and 2010) that mapped the prevalence of the disease in children aged 7–14 for nearly seven decades. We also employed statistical models to investigate which environmental, economic, or demographic factors are associated with the disease in several municipalities. The results showed that the decrease in schistosomiasis from the 1950s to the 2010s was statistically significant, suggesting that improvements in water supply and sanitation conditions require structured and systemic approaches for controlling new forms of disease transmission and outbreak.

## Introduction

Over the decades, several countries have tried to control neglected tropical diseases, including schistosomiasis, by establishing measures to intensify their management. Schistosomiasis is endemic in at least 52 countries [1], affecting approximately 240 million people worldwide. This disease is endemic in ten countries on the American continent. However, only Brazil and Venezuela needed to apply preventive chemotherapy for their population in 2020, including more than 2.2 million school-age children [2]. In addition to chemotherapy, which is not sufficient and accessible to all, the World Health Organization (WHO) recommends several strategies to control and eliminate the disease. These measures include access to safe drinking water, improvements in sanitation, health education, and hygiene, besides environmental and disease control management, even though considering that WaSH interventions (water, sanitation, and hygiene) “are expected to provide modest benefits in limiting *Schistosoma* transmission” [3].

Prevalent in tropical and subtropical areas, especially in poor communities without access to drinking water and adequate sanitary sewage, the disease caused by trematode helminths of the genus *Schistosoma* has epidemiological importance. The epidemiology of the disease is especially relevant in children since the absence of infection in this age group would mean the possible interruption of the transmission. On the other hand, eliminating the disease from the population, including adults, especially workers living in large endemic areas, requires improved household and environmental sanitary conditions.

In Brazil, the epidemiological factors of *Schistosoma mansoni* infection show that social, environmental, and control measures conditions contributed to a reduction in the prevalence rate [4]. Although the relationship between schistosomiasis infection and sanitary conditions has been frequently demonstrated in local or regional studies, nationwide and longitudinal studies can contribute to understanding disease dissemination as well as its explanatory factors throughout the Brazilian territory. Similarly, the approaches to understanding and analyzing schistosomiasis should not be exclusive, and historical, socioeconomic, and cultural elements that determine the disease should also be taken into account [5].

Brazil has extensive experience in conducting prevalence surveys, as demonstrated by surveys covering a wide range of the country and an extended time of approximately seven decades. The first of these surveys was carried out in the 1950s by sanitarians Amílcar Barca Pellon and Isnard Teixeira of the Health Organization Division of Brazil’s Ministry of Health (Divisão de Organização Sanitária do Ministério da Saúde) [6,7]. They examined schoolchildren aged 7–14 through parasitological stool analysis using the spontaneous sedimentation technique based on Hoffman’s technique [8]. In the first phase of the survey, the prevalence was 10.1% for 11 states from the most highly endemic regions, and in the second phase, which surveyed five other states, the prevalence was 0.1%.

Given the epidemiological and social impact of the disease on the population, two decades later, a second national survey of students of the same age group was conducted. The survey was promoted after the establishment of the Special Schistosomiasis Control Program (PECE) (Programa Especial de Controle da Esquistosomose) [9]. This program, which focused on mass drug administration and developed stool testing using the Kato-Katz method [10], reported a disease prevalence of 6.7%.

In recent years, a national survey on the prevalence of schistosomiasis and soil-transmitted helminth infections (INPEG) (Inquérito Nacional de Esquistosomose e Geohelmintose), the first to cover every state in the country, using the Kato-Katz method for stool testing, reported a decrease of the prevalence to 1.79%. Hence, the timeline reveals a reduction in prevalence over the decades [11]. Concomitantly, this survey also reported that the prevalence reached 10.67% in states where the disease is endemic. Moreover, these areas still live with the disease. These data demonstrate that schistosomiasis is still epidemiologically relevant [12] since, from the point of view of the infected patient and public health, there should be no acceptable level of morbidity due to this disease [3]. Hence, this study uses sampling data from a population of schoolchildren aged 7–14 to analyze the behavior of the prevalence of schistosomiasis and the impact on prevalence of access to water and sanitation services. The analysis is based on surveys conducted in Brazilian municipalities over seven decades.

## Methods

### Study design

The epidemiological design of the research consists of a longitudinal study on three panels containing observational ecological data. The outcome variable was the prevalence of schistosomiasis in schoolchildren from seven to 14 years old, arranged by municipality and survey period. The explanatory variables represent population groups arranged by municipality according to the characteristics of ecological studies [13].

### Studied period and data source

Data were extracted from the three national surveys of schistosomiasis prevalence, as follows:

i. The National Helminthological Survey of Schoolchildren (IHE) (Inquérito Helmintológico Escolar) by Pellon & Teixeira, conducted from 1947–1952 in two phases [6,7]. The first phase included 11 states considered endemic for the disease, with a sampling plan that addressed locations of more than 1,500 inhabitants in which 440,786 schoolchildren were examined. In the second phase, locations of more than 1,250 inhabitants of five non-endemic states were included, and 174,192 schoolchildren were examined. In both phases, all regions of the country were sampled, except for the North region, besides municipal districts deemed most economically important. These municipalities represent areas or territories corresponding to municipalities, villages, districts, or regions with nuclei of more than 250 households. In this way, 1,190 locations were surveyed, totaling 614,978 students examined.
ii. PECE [9], conducted from 1975–1979. This survey consisted of a non-probabilistic sample of 327 municipalities in 18 states and areas that were disease-free or endemic, in which 447,779 schoolchildren aged 7–14 were examined. Nevertheless, there is no official information on the criteria used to select the sampled schools for this survey.
iii. INPEG [11], conducted from 2011–2015. This survey also considered schoolchildren aged 7–14 by applying a cluster sampling plan, with areas categorized in three endemic levels (municipalities in non-endemic, low prevalence, and high prevalence areas) and four categories of population size (fewer than 20,000, between 20,000 and 150,000, between 150,000 and 500,000, and more than 500,000 inhabitants). Thus, samples were drawn from those categories to determine the analyzed municipalities, elementary schools, and school classes. As a result, 197,564 examinations of schoolchildren from 521 municipalities representing all Brazilian states were conducted.

The broad extension of the Brazilian territory affected the implementation time of each of the three surveys. Therefore, the impossibility of collecting data in just one year led to the need for around five years of data gathering for each survey. Comparing diagnostic methods between the surveys, although the first used spontaneous sedimentation in the water, the Kato-Katz method applied in the last two surveys has been considered of superior sensitivity since the 1970s. In addition, the Kato-Katz method is currently the gold standard method recommended by the WHO and the Ministry of Health [3,14]. Another difference among surveys is the students’ age group considered. All surveys considered schoolchildren aged 7–14, but just the last one, conducted from 2011–2015, also included students aged 15–17. This inclusion resulted from the eventual presence of some students aged 15–17 in the school classes drawn for examination at the time of the survey. However, according to the methodology presented by INPEG, there was no significant difference in prevalence with the inclusion of these students [11]. Figure 1 describes the surveys, including their respective sampling strategies. Supplementary File S1 Text provides additional details on each survey’s characteristics and specific features.

**Fig 1:**
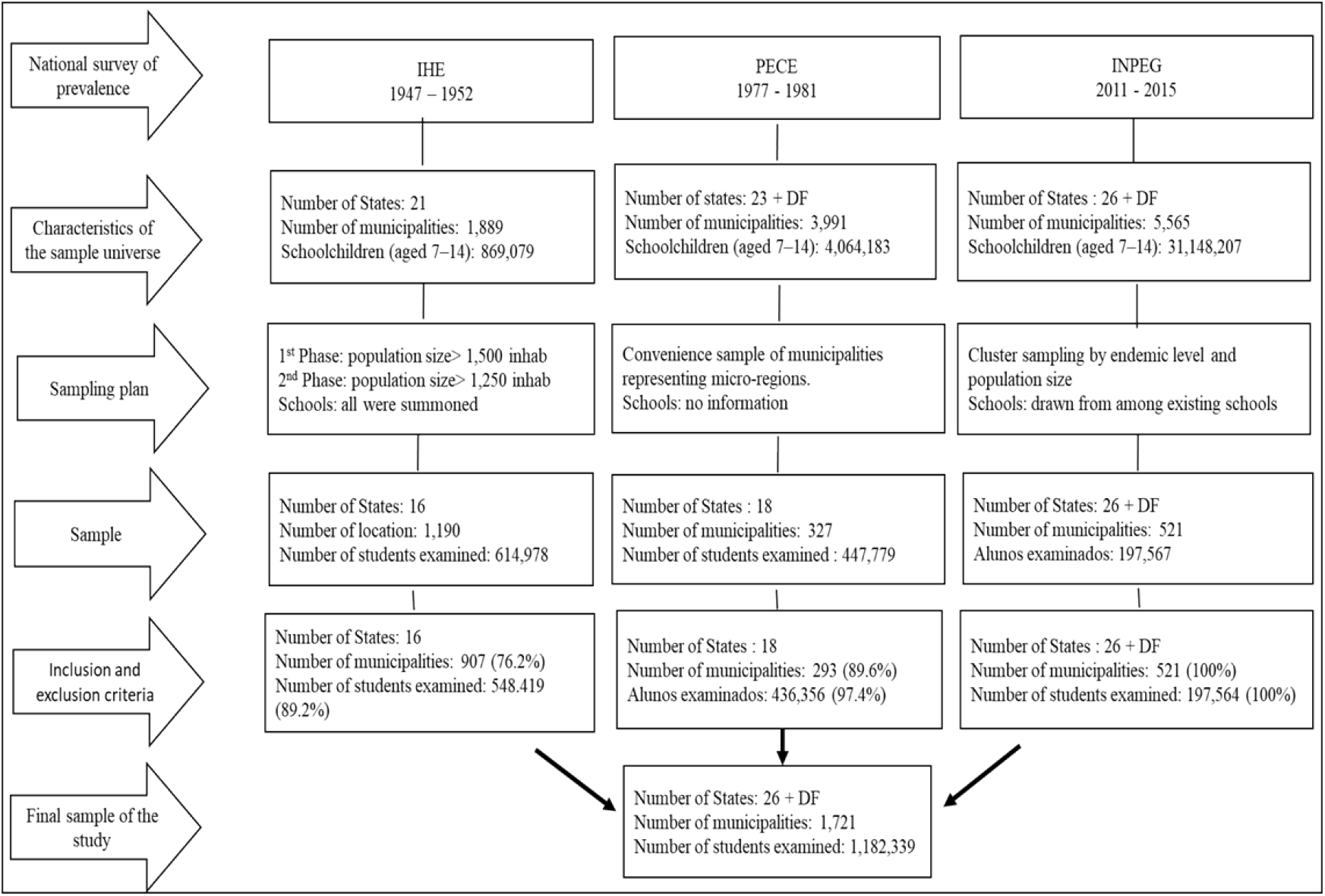
Descriptive flowchart of the three national surveys on the prevalence of schistosomiasis mansoni in Brazil. IHE: National Helminthological Survey of Schoolchildren. PECE: Special Schistosomiasis Control Program. INPEG: National Survey of Prevalence of Schistosomiasis and Soil-transmitted helminth infections. DF: Federal District.

Data related to the explanatory variables were collected from the 1950, 1960, 1970, 1980, 2000, and 2010 demographic censuses for intercensal estimates of the Brazilian Institute of Geography and Statistics (Instituto Brasileiro de Geografia e Estatística – IBGE) and the Institute of Applied Economic Research (Instituto de Pesquisa Econômica Aplicada – IPEADATA) (Table 1).

**Table 1:** Description of evaluated outcome and explanatory variables, periods, and data source.

| Variable | Description | Source | Period |  |  |
| --- | --- | --- | --- | --- | --- |
|  |  |  | IHE | PECE | INPEG |
| Prevalence of schistosomiasis | Number of students with positive stool tests / Total number of examined students | National surveys | 1947–1952<sup>a</sup> | 1975–1979<sup>a</sup> | 2011–2018<sup>a</sup> |
| % of water supply | Number of dwellings with internal piped water supply from the general distribution network / Total number of dwellings | IBGE Census | 1950 | 1970 and 1980<sup>b</sup> | 2000–2010<sup>c</sup> |
| % of sanitary sewage | Number of dwellings with sanitary facilities with drainage connected to the general sewage networks / Total number of dwellings | IBGE Census | 1950 and 1960<sup>d</sup> | 1970–1980<sup>b</sup> | 2000–2010<sup>c</sup> |
| % of urbanization | Number of inhabitants in the urban area / Total number of inhabitants | IBGE Census | 1950 | 1970 and 1980<sup>b</sup> | 2000–2010<sup>c</sup> |
| % literacy rate | Number of people aged 15 years old or older / Total population of the same age group | IBGE Census | 1950 | 1970–1980<sup>b</sup> | 2000–2010<sup>c</sup> |
| % Occupancy condition of households | Number of permanent households in occupancy and owning conditions / Total number of permanent households | IBGE Census | 1950 | 1970–1980<sup>b</sup> | 2000–2010<sup>c</sup> |
| Municipal GDP per capita | Municipal GDP at constant prices – R\$/ total population in the municipality | IPEADATA | 1949 and 1959<sup>b</sup> | 1970–1975–1980<sup>b</sup> | 1999–2010<sup>c</sup> |
| Period | Variable with three categories corresponding to the periods of the surveys | – | 1950 (reference) | 1977 | 2013 |

<sup>a</sup>: Year interval used to define 1950, 1977, and 2013 midpoints for the collection and treatment of explanatory variables. <sup>b</sup>: explanatory variables calculated by applying linear interpolation techniques. <sup>c</sup>: explanatory variables calculated using linear and polynomial interpolation and extrapolation techniques. <sup>d</sup>: values calculated by the population trend method or Apportionment Method (AiBi projection) [15]. National Helminthological Survey of Schoolchildren (Inquérito Helmintológico Escolar – IHE). National Survey on the Prevalence of Schistosomiasis and Soil-transmitted helminth infections (INPEG). Brazilian Institute of Geography and Statistics (IBGE). Institute for Applied Economic Research (IPEADATA). Ministry of Health (MS). Special Schistosomiasis Control Program (PECE). Gross Domestic Product (GDP).

### Inclusion and exclusion criteria

The intense evolution of the political-administrative organization of Brazilian states and municipalities over the decades, reflected in the number of currently existing municipalities, led to the adoption of inclusion and exclusion criteria for this study. The inclusion criteria were defined as: (i) sampled municipalities with territorial delimitation compatible with the demographic censuses of each analyzed period; (ii) sampled municipalities and/or municipal districts that, even incorporated to or emancipated from other municipalities or districts during the period of the three surveys (1947–1953, 1975–1979 and 2011–2015), had available legislative and historical information on their establishment or division process; (iii) sampled municipalities that met the criterion of quality of registration. Subsequently, according to the assumed criteria, the municipalities in which it was not possible to detail the evolution of their establishment, fusion, or incorporation, as well as those lacking enough records for the explanatory variables, were excluded. Figure 1 and Supplementary Note S2 show the complete description of the methodological inclusion and exclusion criteria.

After applying the inclusion and exclusion criteria, the remaining 1,721 municipalities in the study, per survey period, were: 907 for 1947–1953, 293 for 1975–1979, and 521 for 2011–2015. The number of repeated municipalities considering all combinations of survey periods was: 161 for 1947–1953 and 1975–1979, 57 for 1975–1979 and 2011–2013, 146 for 1947–1953 and 2011–2013, and just 41 municipalities for the three surveys together. The survey of 1950 included 16 units in both phases, followed by the 1977 survey with 18, while 26 federal units plus the Federal District were included in the survey of 2013. All regions of the country were sampled in the three surveys, except for the North region in 1950, due to its not yet constituted political-administrative delimitation.

### Outcome variable

The outcome variable of the study was the prevalence of *infection with Schistosoma mansoni* in samples of schoolchildren from seven to 14 years old per municipality (Table 1).

### Independent variables

The independent (exploratory) variables consisted of coverage of water supply and sewerage, and municipalities’ sociodemographic and socioeconomic variables, such as population size, percentages of urbanization and literacy, *per capita* gross domestic product, and the survey period. These variables were defined considering the factors related to infection as indicated in the literature and the context and availability of data in the information systems for each period, seeking to adjust the uniformity and consistency over time. Relevant factors related to the disease, such as family income, coverage of deworming treatment, water treatment for inactivating human schistosome cercariae or chemical molluscicide treatment, malacological surveys, and family or school hygiene practices, could not be included since there are not enough available data from all studied municipalities in the different periods, mainly the 1950s and 1970s.

Since data on the prevalence of schistosomiasis in each survey refer to annual intervals, the midpoint of each interval was adopted as a reference period for collecting and treating explanatory variables. Thus, 1950, 1977, and 2013 were considered these references. Projection and/or interpolation techniques were used in cases where information about the explanatory variables was not available in the reference year. For 1950, we applied a projection by the population trend method — AiBi projection or Apportionment Method — for the sanitary sewage [15]; and a projection by interpolation for the municipal gross domestic product (GDP) *per capita* using data from 1949 and 1959. Projection for sanitary sewage is justified because there is no municipal information on the presence of a sewage collection network since the census of 1950 only reported the presence (or absence) of a toilet at the residence. That makes it difficult to harmonize and standardize this variable in the three analyzed periods; since information about sanitary sewage was available at the state level, the AiBi (or projection) was used to create such an information at the municipal level. As for municipal GDP per capita, the projection is justified because no information was available for the year corresponding to the 1950 reference period. For explanatory variables in the 1977 period, estimates were made using linear interpolation techniques. For the period of 2013, interpolation estimates were performed for 2000 and 2010, and then we extrapolated linear and geometric growth for 2011, 2012, and 2013. The projections for the years 1977 and 2013 were adopted because the Brazilian census information is collected every decade, therefore, in a non-annual series [16,17].

### Data analysis

Since the national surveys are not continuous, the sampled municipalities were not the same for all periods, resulting in an unbalanced data panel with different municipalities in each sampling period. Thus, it was possible to conduct a longitudinal study from three cross-sectional studies to evaluate trends in the prevalence rates of the infection over time and their associations with economic, health, and social indicators for a total of 1,721 municipalities sampled during the periods represented by the reference years 1950, 1977, and 2015.

Descriptive analyses were performed for the municipal data in each period. In the inferential analyses, multilevel statistical models were fitted to estimate the prevalence of schistosomiasis, considering data from the 1,721 sampled municipalities. According to the official territorial division proposed by the IBGE, the Brazilian political-administrative organization is divided into five macro-regions, which include 26 federal units (states) plus the Federal District and 5,570 municipalities. In order to consider this hierarchical characteristic of the data, we applied Generalized Mixed Linear Models (GLMMs) with random effects related to three levels: regions (level 1), states (level 2), and municipalities (level 3). These three hierarchical levels of data were incorporated into the random intercepts of the GLMMs to allow the joint modeling of data from the different municipalities of each sampling period.

The GLMMs also included fixed effects related to the independent variables previously described. We considered the Poisson and Negative Binomial distributions in the analyses, with and without zero inflation for both cases[18–20]. Thus, we allowed the modeling of different data characteristics, such as overdispersion and zero inflation.

A backward selection procedure was used to identify the significant fixed effects, considering a 25% significance level for the removal of an explanatory variable. Thus, at each step of the analysis, the explanatory variable with the highest p-value, among those with a p-value>0.25, was removed from the model. Upon selection of the final models, all variables maintained were significant at the 5% significance level.

The final regression model for each case (Poisson or Negative Binomial distribution, with or without zero inflation) was chosen according to the following parameters: (i) lowest value for the Akaike Information Criterion (AIC); (ii) lowest value for the Bayesian Information Criterion (BIC); and (iii) lowest residual variance [21].

The software EPI INFO 7.1.1 and Microsoft Office Excel 2010 were used for database construction. Descriptive and inferential analyses were performed in the *software R* (version 3.0.2) 2013 (The R Foundation for Statistical Computing) using the statistical packages *lme4* and *glmmTMB*.

## Results

### Descriptive analysis

Table 2 shows the results of the descriptive analysis for the prevalence of schistosomiasis. Despite the large amplitudes, the mean prevalence of infection decreased between the three analyzed periods, with 8.3% for reference period 1950 (SD 17.2), 4.8% for 1977 (SD 12.4), and 0.8% for 2013 (SD 3.5). In addition, the median and amplitude of prevalence were 0.2 and 90.9 in 1950; 0.0 and 71.2 in 1977; and 0.0 and 50.0 in 2013.

**Table 2:** Descriptive statistics on the prevalence of schistosomiasis and independent variables per study period in the 1,721 sampled Brazilian municipalities.

|  | 1947–1953 (n = 907) |  |  |  | 1975–1979 (n = 293) |  |  |  | 2011–2015 (n = 521) |  |  |  |
| --- | --- | --- | --- | --- | --- | --- | --- | --- | --- | --- | --- | --- |
| **Dependent variable** | Mean | SD | Median | AMP | Mean | SD | Median | AMP | Mean | SD | Median | AMP |
| Prevalence of schistosomiasis | 8.3 | 17.2 | 0.2 | 90.9 | 4.8 | 12.4 | 0.0 | 71.2 | 0.8 | 3.5 | 0.0 | 50.0 |
| **Independent variables** | Mean | SD | Median | AMP | Mean | SD | Median | AMP | Mean | SD | Median | AMP |
| Urbanization | 25.6 | 17.4 | 20.6 | 97.0 | 47.4 | 24.5 | 41.6 | 96.6 | 68.4 | 23.2 | 69.1 | 86.3 |
| Literacy | 38.6 | 15.5 | 36.8 | 77.7 | 59.8 | 17.3 | 60.6 | 76.3 | 84.1 | 10.0 | 85.7 | 88.3 |
| Water supply | 6.5 | 10.4 | 1.5 | 73.0 | 30.0 | 22.0 | 24.9 | 90.7 | 71.6 | 21.4 | 75.2 | 100.0 |
| Sewerage | 2.6 | 4.7 | 0.0 | 28.8 | 8.6 | 16.2 | 0.0 | 73.1 | 30.6 | 30.8 | 20.5 | 98.7 |
| % Occupancy condition of the households | 54.9 | 21.1 | 55.4 | 91.5 | 66.2 | 15.1 | 66.3 | 85.4 | 76.3 | 9.1 | 76.7 | 51.9 |
| Municipal GDP per capita | 0.9 | 0.7 | 0.7 | 6.0 | 2.9 | 2.4 | 2.2 | 13.6 | 5.8 | 5.6 | 4.2 | 49.2 |

AMP: Amplitude as maximum value – minimum value. SD: Standard Deviation. GDP: gross domestic product at market price in Brazilian Reais (BRL) per year.

Table 2 also shows descriptive statistics for the explanatory variables that composed the study. They all showed increasing values between 1950 and 2013, especially the sanitary variables related to water supply and sewerage coverages. On average, urbanization went from 25.6% to 68.4% (a 2.6-fold increase); literacy from 38.6% to 84.1% (2.1-fold increase); coverage of water supply network from 6.5% to 71.6% (an 11-fold increase); coverage of sewerage from 2.6% to 31.0% (an 11.9-fold increase); condition of occupancy conditions of households from 54.9% to 76.3% (a 1.4-fold increase); and GDP from 0.91 to 5.77 (BRL) (a 6.3-fold increase).

Table 3 shows the hierarchical (multilevel) description adopted in the study, detailing the distribution of the number of municipalities according to regions and federative units in each analyzed period. Regarding the distribution of the studied municipalities along the five geographical regions of the country, 758 (44.0%) belonged to the Northeast, 506 (29.4%) to the Southeast, 206 (11.9%) to the South, 153 (8.9%) to the Central-West, and 98 (5.7%) to the North region. The Northeast and Southeast regions had the highest percentages of municipalities in each survey. In 1950 (n = 907), the survey included 418 (46.1%) municipalities from the Northeast, 317 (34.9%) from the Southeast, 103 (11.4%) from the South, 69 (7.6%) from the Central-West, and no samples from the North region. For 1977 (n = 293), 114 (38.9%) sampled municipalities belonged to the Northeast region, 73 (24.9%) to the Southeast, 50 (17.0%) to the South, 40 (13.7%) to the Central-west, and 16 (5.5%) to the North. Finally, in 2013 (n = 521), 226 (43.4%) municipalities were in the Northeast, 116 (22.3%) in the Southeast, 82 (15.7%) in the North, 53 (10.2%) in the South, and 44 (8.4%) in the Central-West regions.

**Table 3:** Hierarchical levels and the distribution of the 1,721 sampled Brazilian municipalities included in the study according to federative unit (State) and region for each period.

| Level 3 | Level 2 | Level 1 |  |  |  |  |  |
| --- | --- | --- | --- | --- | --- | --- | --- |
| Region | State | Number of municipalities |  |  |  |  |  |
|  |  | IHE (1947–1952) |  | PECE (1975–1979) |  | INPEG (2011–2015) |  |
|  |  | n | (%) | n | (%) | n | (%) |
| Northeast | Alagoas | 21 | 2.32 | 10 | 3.41 | 24 | 4.61 |
|  | Bahia | 123 | 13.56 | NA | – | 47 | 9.02 |
|  | Ceará | 60 | 6.62 | 22 | 7.51 | 21 | 4.03 |
|  | Maranhão | 29 | 3.20 | 16 | 5.46 | 23 | 4.41 |
|  | Paraíba | 36 | 3.97 | 17 | 5.80 | 21 | 4.03 |
|  | Pernambuco | 61 | 6.73 | 13 | 4.44 | 29 | 5.57 |
|  | Piauí | 16 | 1.76 | 12 | 4.10 | 19 | 3.65 |
|  | Rio Grande do Norte | 41 | 4.52 | 13 | 4.44 | 20 | 3.84 |
|  | Sergipe | 31 | 3.42 | 11 | 3.75 | 22 | 4.22 |
|  | Subtotal | 418 | 46–08 | 144 | 38.90 | 226 | 43.37 |
| North | Acre | NA | – | NA | – | 10 | 1.92 |
|  | Amapá | NA | – | NA | – | 5 | 0.96 |
|  | Amazonas | NA | – | NA | – | 15 | 2.88 |
|  | Pará | NA | – | 16 | 5.46 | 19 | 3.65 |
|  | Rondônia | NA | – | NA | – | 13 | 2.50 |
|  | Roraima | NA | – | NA | – | 7 | 1.34 |
|  | Tocantins | NA | – | NA | – | 13 | 2.50 |
|  | Subtotal | 0 | 0 | 16 | 5.46 | 82 | 15.73 |
| Midwest | Distrito Federal | NA | – | NA | – | 1 | 0.19 |
|  | Goiás | 49 | 5.40 | 25 | 8.53 | 18 | 3.45 |
|  | Mato Grosso | 20 | 2.21 | 7 | 2.39 | 12 | 2.30 |
|  | Mato Grosso do Sul | NA | – | 8 | 2.73 | 13 | 2.50 |
|  | Subtotal | 69 | 7.60 | 40 | 13.65 | 44 | 8.44 |
| Southeast | Espírito Santo | 18 | 1.98 | 10 | 3.41 | 16 | 3.07 |
|  | Minas Gerais | 250 | 27.56 | 52 | 17.75 | 56 | 10.75 |
|  | Rio de Janeiro | 49 | 5.40 | 11 | 3.75 | 21 | 4.03 |
|  | São Paulo | NA | – | NA | – | 23 | 4.41 |
|  | Subtotal | 317 | 34.95 | 73 | 24.91 | 116 | 22.26 |
| South | Paraná | 58 | 6.39 | 7 | 2.39 | 21 | 4.03 |
|  | Rio Grande do Sul | NA | – | 25 | 8.53 | 14 | 2.69 |
|  | Santa Catarina | 45 | 4.96 | 18 | 6.14 | 18 | 3.45 |
|  | Subtotal | 103 | 11.35 | 50 | 17.06 | 53 | 10.17 |
| Total |  | 907 |  | 293 |  | 521 |  |

NA: not analyzed. School Helminthological Survey (IHE). National Survey on the Prevalence of Schistosomiasis and Soil-transmitted helminth infections (INPEG). Special Schistosomiasis Control Program (PECE).

### Statistical models

Considering the criteria for comparing the models, the Poisson model with zero inflation presented the best performance, with lower values of residual variance, AIC, and BIC. Table 4 shows the Rate Ratio (RR) estimates for schistosomiasis infection, and the respective 95% confidence intervals (CI) obtained from the zero-inflated Poisson multilevel regression model.

**Table 4:** Results from the zero-inflated Poisson multilevel regression model fitted to assess the prevalence of schistosomiasis mansoni in the sampled Brazilian schoolchildren.

| Variable | Poisson with zero-inflation |  |  |  |
| --- | --- | --- | --- | --- |
|  | RR | (CI 95%) | Estimate | P-value |
| LN Population | 0.862 | (0.825 – 0.901) | -0,148 | <0.001 |
| % Urbanization | 0.991 | (0.989 – 0.993) | -0.009 | <0.001 |
| % Occupancy condition of the household | 0.986 | (0.983 – 0.989) | -0.014 | <0.001 |
| % Water supply | 1.010 | (1.008 – 1.011) | 0.010 | <0.001 |
| % Sewerage | 0.996 | (0.994 – 0.998) | -0.004 | <0.001 |
| Year: 1977 | 1.352 | (1.256 – 1.454) | 0.301 | <0.001 |
| Year: 2013 | 0.214 | (0.184 – 0.249) | -1.542 | <0.001 |
| Model constant (intercept) | 0.004 | (0.001 – 0.027) | -5.488 | <0.001 |

Reference year: AIC: 11162.2. BIC: 11233.1. CI: Confidence interval. Standard deviation of residuals: 65.4. LN: natural logarithm. Residual variance: 4283.5. RR: Rate Ratio.

The explanatory variables that remained in the model of the prevalence of schistosomiasis were the natural logarithm of population size, % urbanization, % occupancy condition of the domicile, % water supply, % sewerage, and the categorical variable related to the survey period (the 1950 period was used as a reference for the analysis).

A negative value in the estimate of the effect of a variable indicates that an increase in its value results in a decrease in the prevalence of infection. This was the case for the variables natural logarithm of population size (−0.148; p-value <0.001), % urbanization (−0.009; p-value <0.001), % occupancy condition of the households (−0.014; p-value <0.001), and % sewerage (−0.004; p-value 0.001). Based on the associated RR, the increase of one unit in the numerical value of these variables causes a decrease of 13.8%, 0.9%, 1.4%, and 0.4% in the estimated mean for the prevalence, respectively. On the other hand, the results showed an inverse effect on the prevalence of infection for the water supply variable, with a positive value for its estimated effect (0.010; p-value <0.001). Concerning the categorical variable representing the survey periods, an estimated increase of 32.5% in the mean prevalence was determined from 1950 to 1977 (0.301; p-value <0.001). However, a significant effect of decreasing prevalence was estimated for 1950–2013 (−1.542; p-value <0.001), representing an average decrease of 78.6% between the first and the last analyzed periods. Based on these results, a significant effect of decreasing prevalence between 1977 and 2013 (−1.843; p-value <0.001) was also estimated, representing an average decrease of 84.2% between these two study periods.

### Ethics statement

This study was conducted exclusively with secondary and aggregated data, publicly accessible and in accordance with resolutions of the National Health Council No. 466/2012 [22] and No. 510/2016 [23], exempt from evaluation by the Research Ethics Committee.

## Discussion

The analysis identified significant effects of environmental, economic, and demographic factors on the prevalence of schistosomiasis by evaluating its trend during the three national surveys. Hence, the study proves for the first time these factors’ effects by conducting an inferential analysis. It is noteworthy that, although the descriptive analysis indicated a decrease in prevalence between 1950 and 1977, the fitted statistical model indicated a significant increase in the mean prevalence for the same period. This outcome may be influenced by the behavior of the other variables included in the model, although it may also result from the combination with other factors. For example, the characteristics of the sampling procedures and/or the possible emergence of new transmission areas could increase positivity or cause a small variation in the percentage of reduction in some regions. This situation was previously observed in two epidemiological studies using data from 1950, 1980, and 1990 in endemic states of the country, indicating that the basic pattern of spatial distribution was similar between the evaluated years and demonstrating the heterogeneity of the historical process of unequal health patterns in the country[24,25].

The results of the statistical models of this study showed that the environmental variables contributed significantly to the prevalence of schistosomiasis. The protective association between the expansion of sewerage coverage and the reduction of prevalence has been portrayed in epidemiological studies since the 1960s[26]. For instance, a significant association between, and high prevalence of schistosomiasis in children with a lack of sewerage or sewage disposal in a cesspool compared to families using a safe sewage network (OR 1.8; CI 1.3 – 2.4; OR 1.9; CI 1.3 – 2.4, respectively) was found [27]. Coincidently, a study by Ximenes et al. (2013) showed that sanitary installation for domestic wastewater collection in septic tanks connected to the general city sewage network was significantly associated with a decreased probability of infection[28]. Kabatereine et al. (2011) also showed that adding an item in the socioeconomic component of the family, such as the latrine, decreased the chances of infection[29]. Even when latrines were available, families’ preference for their use also reduced the occurrence of the disease[30], which was found when households lacked a functional toilet[31,32].

Therefore, although Brazil had sanitary sewage networks in only 60.3% of its municipalities in 2017[33], the impact of this service in interruption of the disease is evident, functioning as a sanitary barrier to fecal contamination in water bodies containing intermediate hosts. The results of this study validate the importance of public policies promoting the implementation of sanitation solutions. According to these results, if municipalities, which have a current coverage of 20% of the sewage system, a common situation in some areas of the country, reach 100% coverage, a 27.4% reduction in the average prevalence of schistosomiasis can be expected, which is an important outcome in terms of public health.

In addition, the treatment and supply of safe drinking water have been considered another environmental variable as an effective and lasting measure to prevent disease[27,34–36]. On the other hand, some studies found no significant association between drinking water supply and reduced prevalence of schistosomiasis [37–40]. Although it is not a waterborne disease, it is associated with water since part of the life cycle of the infectious agent occurs in aquatic snails. Thus, it is reasonable to assume that piped water should not pose a risk of transmission. However, although the results of this study indicate a controversial finding, three possible explanations can be put forward.

Firstly, the infrastructure for piped water supply has expanded over the decades, but this expansion has not guaranteed uninterrupted supply or the quality of water supplied. Even in a more recent period, in 2006, the irregularity in water supply from the public network can affect about 80% of municipalities in certain regions, like the state of Bahia, where schistosomiasis is endemic [41]. Moreover, the Northeast and Southeast regions, which presented the highest prevalence of this disease, exhibited the highest frequencies of systematic interruptions in the water supply in 2020, reaching 66.1% and 46.5%, respectively[42]. This intermittence can lead users to depend on contact with unsafe water sources, contacts that may eventually even be increased at day times of high schistosomiasis transmission. Consequently, even in municipalities having households supplied with piped water, there could be a high probability of infection by the disease. Intermittent water supply can disrupt family dynamics, a situation directly related to obligations that often still fall on women. In a society and economy marked by the sexual division of labor, this dynamic leads to the penalization mainly of women and their children, who end up accompanying their mothers [43] in using unsafe water sources, a risk factor in the dynamics of schistosomiasis transmission, reported since the 1980s[44]. In addition, discontinuity of water supply produces other adverse effects, such as deterioration of water networks designed for continuous supply, leading to leaks and modification of the water quality standard. Consequently, users adapt to meet adversities, highlighting the inequality and vulnerability to shortages to which a city or region is exposed[45]. An intervention study that considered households before and after the installation of piped water showed that the replacement of surface water by piped water utilization occurs significantly only when more water is used (more than 1,000 liters of piped water per person per year). Even more, the use of water bodies, such as rivers and streams, continued after the shutdown of the piped water network due to system failures and the preference for using these natural sources for domestic activities, such as washing clothes[46].

The second explanation regarding disease prevalence despite the availability of piped water is related to a supply insufficiency for some households to eliminate other contact forms with surface water for domestic, leisure, behavioral, or labor use, such as fishing and irrigation. Eventually, the presence of piped water supply may free up more time for residents to perform these activities more frequently, increasing the risk of contamination. When disassociated from facilities for other home uses, such as laundry, sink, and shower, piped water supply can contribute to the continuity or increase of the behavior of accessing transmission sites [47,48]. Another aspect related to the water contact practices was demonstrated by a spatial community study verifying that the public water supply could potentially decrease dependence on surface water. However, this relationship was modified by the quality of the water from the sources of public supply, which was considered poor by domestic users[49].

Thirdly, an aspect probably not strongly related to our results, although worthy of analysis, is the effect of the technology used in surface uptake, adduction, and water treatment on dermal contact and survival of infectious forms of the schistosome. Filtration and chlorination are widely used methods for water treatment in conventional and simplified treatment plants all over the country [50,51]. These processes are credited as likely to produce waters free of contamination from cercariae, depending on storage time, exposure temperature, chlorine concentration, or filtration rates, besides the concentration of cercariae itself[52]. However, there are no current guidelines for the specific care related to water treatment and its respective technical and operational infrastructure in endemic schistosomiasis regions, as demonstrated by other systematic reviews [48,52]. Therefore, operational deficiencies such as lack of water treatment have been observed despite Brazil’s enhanced access to water supply networks and infrastructure since the 1940s. In 1948, shortly before implementation of the first survey included in this study, only 9% of municipalities received treated water, a deficiency even more prominent in rural areas[53]. Incomplete water treatment and deficient distribution systems are still a reality[54] since 11.7% of Brazilian municipalities still lacked operative water treatment plants, either conventional or simplified, in 2017[33].

Other conditions different from environmental factors contributed to the decrease in disease prevalence, such as the condition of household occupation, degree of urbanization, and population size. Low socioeconomic status is a known risk factor for diseases caused by parasitic infections such as schistosomiasis[28,55]. In this study, residents’ housing conditions, such as acquired households and owned rather than rented dwellings, were used as a proxy for socioeconomic status. A similar conclusion was obtained in studies in Pakistan, Bangladesh, and Thailand, with families living in rented houses at increased risk of developing infectious diseases or their symptoms, including parasitic bowel diseases, compared with families who owned their housing [56–58]. Thus, the condition of home ownership was associated as a protective factor against disease, demonstrating that socioeconomic structure can produce and condition the distribution of schistosomiasis in the population.

Regarding the degree of urbanization, recent outbreaks of schistosomiasis have been prevalent in urban and peri-urban environments due to unplanned urbanization [59–62]. On the other hand, rapid urbanization has implications for infectious diseases usually described in rural areas and reduces the risk of exposure to infection in previously endemic areas[63]. Thus, the effect on epidemiological patterns of the relationship between demographic events of inter and intra-regional migratory flows with economic cycles of retraction and expansion of agricultural and industrial activities experienced by the country is undeniable. This relationship generated the model of capitalist expansion and economic growth, sometimes excluding but also enabling the last seven decades of educational, sanitary, economic, and infrastructure improvements that also resulted in changes to epidemiological patterns of infectious diseases [64,65]. Therefore, urbanization is assumed as a protective trait against the disease, which could be echoed in the institutional feasibility of increasing and expanding public health and sanitary policies. The establishment of the Brazilian Unified Health System (SUS), including an alternative model focused on the promotion and prevention of health from the decentralization of strategies and programs for the control of schistosomiasis, is an example of these health and sanitary polices [66,67]. Other public policies, which could correspond to the changes that have occurred over the decades, are water and sanitation services, such as the National Sanitation Plan (Plano Nacional de Saneamento – Planasa), established in 1971 and abolished at the end of the following decade, and the current federal basic sanitation policy from Law No. 11.445/2007 and No. 14.026/2020. Although increased access to public services was considered deeply discriminatory in the 1970s regarding demographic and social criteria and currently poses risks concerning the universal access to services and human rights, they were essential instruments for expanding public water supply and sanitary sewage networks in the country[68,69].

In general, the findings of this research show that the reduction in the prevalence of schistosomiasis in Brazil over seven decades can be explained by the combination of community, demographic, socioeconomic, and specific environmental factors. The ecological design of the study, with the municipality as the unit of analysis, avoids including behavioral and other individual variables in the model, likely associated with infection. The mechanism of schistosomiasis transmission is complex and includes several conditioning factors [70]. Thus, disease control depends on preventive measures, such as early diagnosis and timely treatment, health education, surveillance and control of intermediate hosts, and basic sanitation. It is also noteworthy that the Brazilian regions differ in how their governments administer the promotion of disease control policies, especially among states that differ in aspects like location, territorial extension, and environmental and socioeconomic conditions that could interfere with the disease cycle. Coincident with considerations of the systematic review by Grimes et al. (2014), the difference between infection rates among people with and without access to drinking water and sanitary sewage varies greatly, suggesting that the impact of access to water and sanitation is mediated by other social, behavioral, and environmental factors.

## Limitations

Our results are valid only for the observed sample and, therefore, not generalizable. In addition, other limitations must be considered when interpreting these results. The variables were collected in different periods, and such practice of collecting old census data required a process of harmonization between variables to allow comparisons. Another limitation inherent to census data includes the availability of access-restricted information on public service facilities and not the quality and availability of WaSH services. Further exploration of other data is necessary to understand the positive association between the prevalence of schistosomiasis and the availability of drinking water networks, including the effects of supply interruptions and changes in use based on water quality or behavioral and occupational habits. Finally, we cannot make conclusions on the causality of this association due to a limitation of ecological design.

To the best of our knowledge, this is the first international study, especially in the Americas, that has used a longitudinal epidemiological design to analyze data from national prevalence surveys covering seven decades. The results showed that the prevalence of schistosomiasis infection in schoolchildren in Brazilian municipalities decreased significantly over the decades. This decrease in prevalence of infection may be associated with environmental factors, urbanization, and housing conditions, which have improved over the decades. It is noteworthy that the association with water supply should be carefully interpreted and focused on other possible factors not evaluated here, confirming the need for a systemic approach. In addition, sanitary sewage and health education should be widely offered to the population in their homes and other places, such as work or school environments. Other national prevalence surveys and research should be conducted more continuously to monitor disease positivity and its determinants over time.

## Data Availability

All relevant data are within the manuscript and its Supporting Information files.

## Acknowledgments

We thank the task teams responsible for organizing and operationalizing the research field in all surveys. We are immensely grateful to the researcher Prof. Dr. Naftale Katz, from Instituto René Rachou/Fiocruz Minas, who assisted in making this research feasible by guiding us to the source and acquisition of data and for sharing with us his experience in conducting surveys.

## Supporting information captions

S1 Text. General and methodological characteristics of the three surveys National Helminthological Survey of Schoolchildren (IHE) (1947–1953),, Special Schistosomiasis Control Program (PECE) (1975-1979), and National Survey of Schistosomiasis and Geohelminthiasis Prevalence (INPEG) (2011-2015).

S2 Text. Methodological criteria related to the process of creating municipalities, used for inclusion and exclusion from the study.

## DATA AVAILABILITY STATEMENT

The study was conducted using publicly available data in information systems of the Brazilian Institute of Geography and Statistics (https://www.ibge.gov.br/) as reported in the methods section, Institute of Applied Economic Research (IPEADATA) (http://www.ipeadata.gov.br/Default.aspx). The other data are available to researchers, filed at Instituto René Rachou.

### Financing information

This work was carried out with the support of the Coordination for the Improvement of Higher Education Personnel – Brazil (CAPES) – Financing Code 001 (https://www.gov.br/capes/pt-br), which granted financial aid in the form of a scholarship granted to MCSS. The financier did not participate in the study design, data collection and analysis, publication decision or manuscript preparation. Conflict statement: the authors declare that there are no conflicts of interest.

### Author contribuions

Conceptualization: Mariana Cristina Silva Santos, Léo Heller.

Data Curation: Mariana Cristina Silva Santos, Léo Heller.

Formal Analysis: Mariana Cristina Silva Santos, Guilherme Lopes de Oliveira, Sueli

Aparecida Mingoti, Léo Heller.

Funding Acquisition: Léo Heller.

Investigation: Mariana Cristina Silva Santos, Léo Heller.

Methodology: Mariana Cristina Silva Santos, Guilherme Lopes de Oliveira, Sueli Aparecida Mingoti, Léo Heller.

Project Administration: Mariana Cristina Silva Santos, Léo Heller.

Resources: Mariana Cristina Silva Santos, Léo Heller.

Software: Mariana Cristina Silva Santos, Guilherme Lopes de Oliveira, Sueli Aparecida Mingoti.

Supervision: Sueli Aparecida Mingoti, Léo Heller. Validation: Sueli Aparecida Mingoti, Léo Heller.

Visualization: Mariana Cristina Silva Santos, Guilherme Lopes de Oliveira, Sueli Aparecida Mingoti, Léo Heller.

Writing – Original Draft Preparation: Mariana Cristina Silva Santos, Guilherme Lopes de Oliveira, Sueli Aparecida Mingoti, Léo Heller.

Writing – Review & Editing: Mariana Cristina Silva Santos, Guilherme Lopes de Oliveira, Sueli Aparecida Mingoti, Léo Heller.

